# Strain estimation in aortic roots from 4D echocardiographic images using medial modeling and deformable registration

**DOI:** 10.1101/2022.11.04.22281143

**Authors:** Ankush Aggarwal, Peter Mortensen, Jilei Hao, Łukasz Kaczmarczyk, Albert T. Cheung, Lourdes Al Ghofaily, Robert C. Gorman, Nimesh D. Desai, Joseph E. Bavaria, Alison M. Pouch

## Abstract

Even though the central role of mechanics in cardiovascular system is widely recognized, estimating mechanical deformation and strains in-vivo remains an ongoing practical challenge. Herein, we present a semi-automated framework to estimate strains from four-dimensional (4D) echocardiography images and apply it to the aortic roots of patients with normal trileaflet aortic valves (TAV) and congenital bicuspid aortic valves (BAV). The method is based on fully nonlinear shell-based kinematics, which divides the strains into in-plane (shear and dilatational) and out-of-plane components. The results indicate that, even for size-matched non-aneurysmal aortic roots, BAV patients experience larger regional shear strains in their aortic roots. This elevated strains might be a contributing factor to the higher risk of aneurysm development in BAV patients. The proposed framework is openly available and applicable to any tubular structures.

## 1. Introduction

Mechanical strains experienced by cardiovascular tissues are closely related to their function and long-term adaptation [1]. The strains affect the cellular behavior and phenotype, which over time can lead to changes in the tissue microstructure [2]. This is especially relevant for arterial tissues that experience high pulsating pressures and shear stresses from the blood flow, and arterial stiffening is associated with normal aging process [3]. In arteries, excessive strains can also cause damage and tear in the tissue, sometimes creating a false lumen [4, 5]. Thus, tissue strains can carry extra information that improves patients’ risk stratification [6, 7] and could potentially serve as early indicators of multiple vascular diseases, such as aneurysm, dissection, and stenosis. A large number of studies have focused on calculating tissue strains by combining insilico modeling and ex-vivo experimentation [8]. However, using ex-vivo/in-silico models to predict or even inform in-vivo values is far from trivial. Multiple invivo features, such as pre-strain [9], inelastic behavior [10] and moving boundary conditions [11], are extremely challenging to replicate.

A capability to measure tissue strains in-vivo can transform the vascular research landscape, by allowing direct monitoring of these important biomechanical indicators, determining differences between patient subgroups, and informing/validating the in-silico models. Recent advances in four-dimensional (4D) imaging provide an opportunity to make progress in this direction. 4D in-vivo images, such as those acquired using transesophageal echocardiography (TEE), contain an extensive amount of information. Yet, only the 3D tissue and organ shape characteristics are regularly extracted and utilized from these images. In addition to the shape, the tissue motion captured in the 4D images can be further processed to estimate in-vivo strains. While such strain measurements are now being investigated as functional indicators for the cardiac tissue [12], the same is not true for vascular tissues. For example, aneurysm diameter remains the primary feature used by clinicians to decide whether a surgical intervention is required.

Numerous studies have proposed strain estimation using 2D ultrasound (US) imaging by cross-correlation and speckle tracking, with a wide range of applications to the myocardium [13–16], aortic wall [17–19], carotid artery [20–22], prostate [23] and skin [24]. Commonly known as US strain imaging, the underlying approach is to use cross-correlation in the raw signal and/or image domain to determine the tissue velocity and displacement, which is further processed to estimate strains. However, 2D ultrasound has limitations. The out of plane motion may cause signal decorrelation and affect the quality of results [25], and the associated out of plane strains are challenging to estimate. Indeed, many of the studies only report the changes in diameter. Moreover, further processing is required to capture the discontinuities between tissue and lumen [26]. A similar approach is used in Tissue Doppler Imaging [27], which has also been extended to 3D [28, 29].

Multiple studies have reported full 3D strain estimates in ventricles using tagged magnetic resonance imaging [30, 31] and 3D speckle tracking in 4D ultrasound [32–37]. However, these are structure-specific and rely on vendor-specific proprietary software, which have shown suboptimal intervendor agreement of strain measurements [38]. Even though 4D TEE plays a valuable role in aortic pathologies assessment and repair [39, 40], strain estimation in arteries remains a challenge due to their smaller thickness when compared to ventricles. Some have applied the motion tracking designed for ventricles to aortic vessels by adding a fictive pseudo-apex[41–44]. However, these motion tracking algorithms are optimized for ventricles and specific to the image acquisition hardware. In most of these studies, only global strain estimates, such as changes in diameter were estimated. Another approach has been to track the vessel wall motion and couple it with inverse finite element analysis to estimate strains and stresses [45– 47]. However, to the authors’ knowledge, few studies have directly estimated regional in-vivo strains in thin tissues [48].

The aim of the presented work is to fill the gaps identified above with an open-source framework to estimate 3D tissue strains in thin tubular structures, such as arteries, from 4D echocardiographic images. One of the methodological novelties of our framework is the use of a medial model, which conforms well to thin tubular structures and appropriately quantifies in-plane and out-of-plane strains, and at the same time facilitates population-level studies through spatial correspondence between patients. To demonstrate the proposed framework, the aortic root is chosen as an application for reasons explained next.

The aortic root connects the left ventricle to the ascending aorta and supports the cusps of the aortic valve (AV). It plays an important role in several pathologies, such as aortic aneurysm and dissection. Moreover, its biomechanical coupling with the AV is complex. Patients with a bicuspid aortic valve (BAV)

[49] are known to be at higher risk of aortic aneurysm compared to those with a normal, trileaflet AV (TAV). However, the underlying reasons for higher risk of root aneurysm are not fully known. The 4D dynamics of aortic root has been analyzed in detail [50] and differences have been found in the enclosed volume (also known as the lumen volume) for TAV versus BAV patients [51]. However, detailed in-vivo strains cannot be yet quantified from the 4D dynamics. The only information available about the in-vivo strains experienced by the aortic root tissue is through invasive animal studies using physical markers [52–54]. By applying the proposed novel framework, the objective of this work is to provide the first in-vivo strain measurements in human aortic roots and ascertain any differences between TAV and BAV patients. Relative to transthoracic echocardiography (TTE), TEE is superior for aortic root strain assessment given the close proximity of the transesophageal transducer to the valve apparatus and is therefore used in this study. However, the proposed framework is not specific to TEE and could be generalized to other imaging modalities with which high quality images of vasculature can be obtained.

In the next section, details of the proposed framework to estimate tissue strains from TEE are provided. Given the thin profile of aortic root, strains are divided into in-plane and out-of-plane using shell kinematics [55–57]. Moreover, to account for the large deformations that the aortic root undergoes during the cardiac cycle, fully nonlinear kinematics are used. In Section 3, the results for size-matched TAV and BAV aortic roots are compared in terms of reference shape, global strains, and regional strains. In Section 4, we discuss the implications of the results and possible extensions of the proposed method in the future.

## 2. Methods

### 2.1. Image acquisition

Electrocardiographically gated real-time 3D TEE images of the aortic root were retrospectively acquired from 35 patients with the approval of the Institutional Review Board at the University of Pennsylvania. The patients included 16 with physiologically normal TAVs and ascending aortas and 19 with minimally calcified BAVs with or without aneurysm of the aortic root and/or ascending aorta. The 3D TEE images were obtained using the iE33 imaging platform with the X7-2t xMATRIX transducer (Philips Medical Systems, Andover, MA) at end-expiration during positive pressure ventilation in anesthetized patients to eliminate motion caused by respiration. To compare the roots with similar sizes, only the roots with a maximum radius^1^ between 15.5 mm and 18 mm were included, which resulted in a dataset of 10 TAV and 7 BAV roots. Each patient’s 3D TEE image series consisted of 8 to 40 frames showing the aortic root from the level of the left ventricular outlet (LVO) to the sinotubular junction (STJ) over one complete cardiac cycle beginning at early systole. The images were exported in Cartesian DICOM format with nearly isotropic voxel size ranging from 0.4 to 0.8 mm.

### 2.2. Image segmentation

From each patient’s 3D TEE dataset, a series of 3D aortic root reconstructions was generated over one cardiac cycle using the custom semi-automated image analysis pipeline illustrated in Figure 1. Briefly, the aortic root was first manually traced in a systolic frame of the cardiac cycle using ITK-SNAP, an open-source tool for interactive 3D medical image segmentation[58]. The segmentation was annotated at the STJ, LVO, and interatrial septum to define the orientation of the root for subsequent measurement. This segmentation was referred to as the “reference segmentation” and was verified by a second observer. A medial model was automatically created from the segmentation as described in Section 2.3. Next, the reference segmentation was morphologically dilated to define a region of interest around the aortic root in the image. Then the TEE image series was down-sampled, and deformable registration between each consecutive pair of 3D volumes was performed. The resulting deformation fields were used to propagate the reference mask to all 3D volumes in the series. The reference frame was then registered at full resolution to all other 3D volumes in the TEE series using the reference masks in each frame for increased computational efficiency and precision. The medial model of the reference frame was then propagated to all other frames, producing a series of 3D medial models of the aortic root at all available frames in the cardiac cycle. Note that image down-sampling in the first registration step was performed only to quickly define a region of interest around the root in each 3D frame, whereas full-resolution deformable registration was guided by these regions of interest and used to propagate the aortic root model to all available 3D frames. All deformable registrations were performed using the generalized open-source greedy 3D registration tool [59]. The same Gaussian regularization parameters were used for all data sets: metric gradient smoothing with a sigma value of 3mm and deformation field smoothing with a sigma value of 1.5mm.

**Figure 1:**
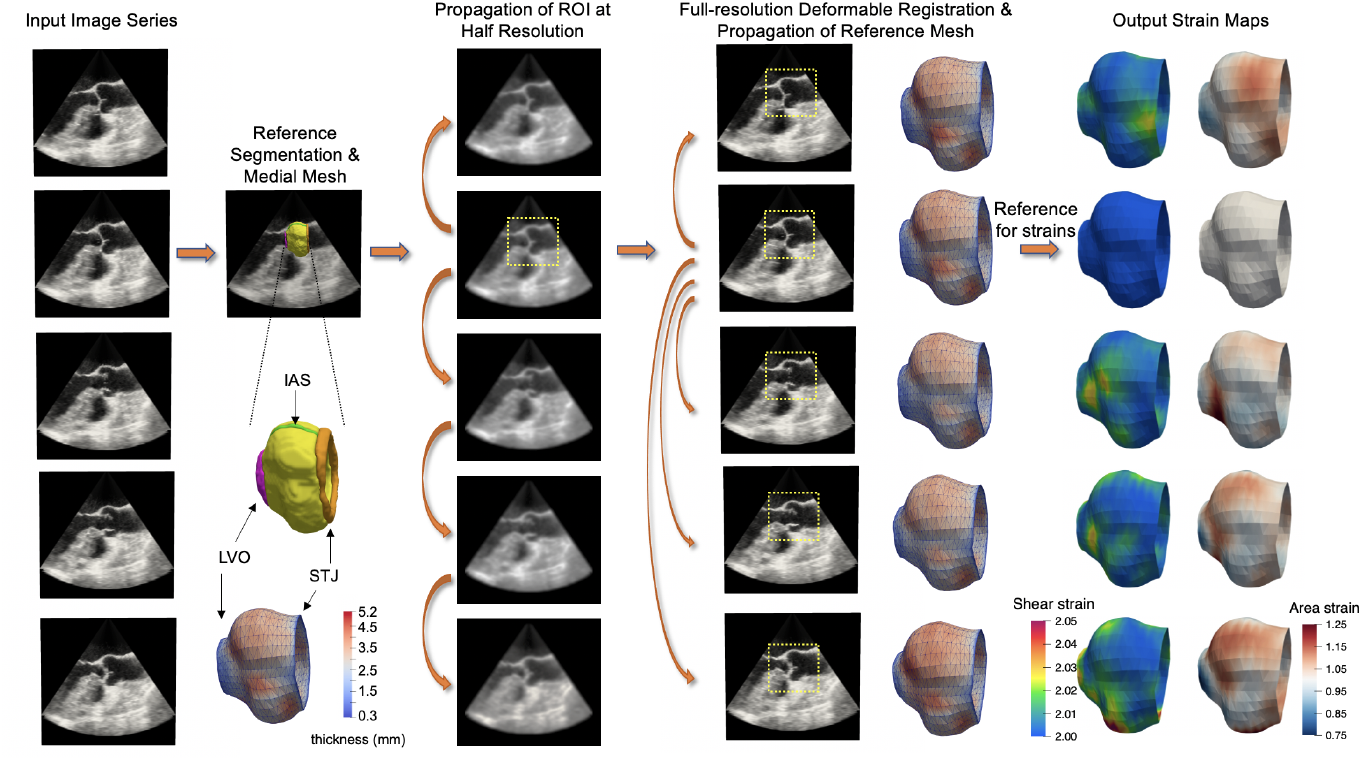
The proposed semi-automated image segmentation, modeling, and strain analysis pipeline. A reference 3D volume is selected from the input image series and manually segmented. The aortic root is labeled in yellow, sinotubular junction (STJ) in orange, left ventricular outlet (LVO) in pink, and interatrial septum (IAS) in green. A medial mesh, with thickness shown in color, is generated from the reference segmentation. Consecutive pairs of images are then registered at half-resolution in order to propagate a masked region of the root to all frames in the series. Next, full-resolution deformable registration is performed between the reference frame and all other frames in order to propagate the reference medial model to all time points in the series. Maps of shear and area (dilatational) strain are computed from the series of medial models of the aortic root.

### 2.3. Medial modeling

The shape of the aortic root is modeled at each cardiac phase using medial axis representation in a manner that preserves spatial correspondences across time and between patients. The medial axis, or morphological skeleton, of an object is a surface formed by the centers of all maximally inscribed balls (MIBs) in the object [60], as illustrated in 2D in Figure 2. The medial axis can be parameterized by (**m**, *R*) ∈ ℝ^3^ × ℝ where **m** is a medial surface formed by the centers of all MIBs (or maximally inscribed disks in 2D) in an object and *R* is a scalar radial thickness field defined over the medial surface. *R* refers to both the radius of a MIB centered on the medial axis and, equivalently, to the distance between the medial axis and object boundary. Any point **m** on the interior of the non-branching medial axis is associated with two spokes (vectors) pointing to two points, *b*^+^ and *b*^*−*^, on the object boundary. In Figure 2, the points *b*_1_^+^ and *b*_1_^*−*^ are said to be medially linked, because they are points of tangency of the MIB centered at the point **m**_1_ on the medial axis. The point *b*_0_ is not medially linked to any other boundary point because it is a point of tangency of a disk centered at an edge of the medial axis. Since the thickness of an object is related to the scalar radius function, medial axis representation is useful for quantifying locally varying thickness of sheet-like anatomical structures. In this work, each point on the medial axis is associated with a measurement of localized thickness of the aortic root, defined as the distance between medially linked boundary points (e.g., the distance between *b*_1_^+^ and *b*_1_^*−*^ in Figure 2).

**Figure 2:**
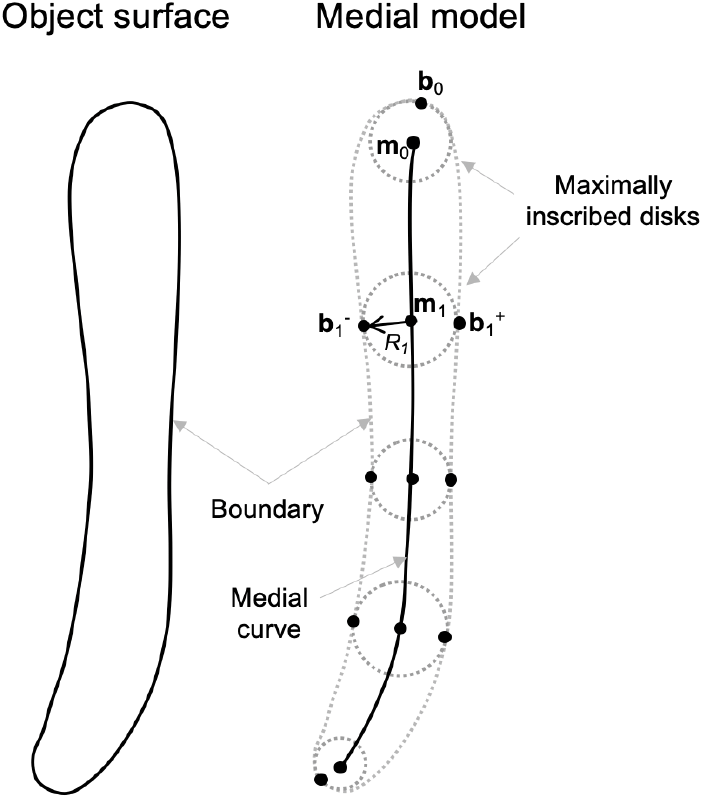
The 2D medial geometry of an thin oblong object that has a non-branching medial axis. The point **m**_0_ is the center of a maximally inscribed disk that is tangent to the object boundary at *b*_0_. The maximally inscribed disk centered at **m**_1_ has radius *R*_1_ and is tangent to the object surface at two medially linked boundary points: *b*_1_^+^ and *b*_1_^*−*^.

Given a segmentation of an anatomical structure, its medial axis can be modeled using several strategies. For example, deterministic skeletonization algorithms estimate an object’s medial geometry directly from its boundary representation (e.g., [61–63]), while inverse skeletonization algorithms approximate an object’s medial geometry by fitting a pre-defined deformable model with fixed medial topology to the object (e.g., [64, 65]). In this work, we use a boundary contour resampling scheme adapted from [66] to generate a 3D triangulated medial mesh of the aortic root based on the geometry of its boundary (similar to deterministic algorithms) while assuming a non-branching medial axis topology (similar to inverse skeletonization approaches). The meshing algorithm is specific to 3D shapes with non-branching medial geometry, meaning that the 3D shape can be sliced into 2D cross-sections that are oblong and homeomorphic to a disk (e.g., placentas, cardiac ventricles, kidneys). The input to the algorithm is a segmentation or boundary representation of an anatomical structure, and the output is a triangulated mesh that approximates the medial surface of the structure.

An overview of the 3D medial meshing process is illustrated in Figure 3. The 3D aortic root shape is first rigidly transformed so that the aortic outflow tract is aligned with the z-axis: the STJ is up, the LVO is down, and the interatrial septum is aligned with the x-axis. The segmentation is then cross-sectioned into long-axis slices that are oblong and homeomorphic to a disk (i.e., they have a closed, flat geometry that can be approximated by a non-branching medial axis representation). For each longitudinal 2D cross-section of the root, a binary image is created, morphologically closed, and smoothed, as shown in Figure 3C. Let *B*(*γ*) : [0, *M*) → ℝ^2^ be a closed curve parameterized by arc length that outlines the boundary of the root and is computed from the zero level-set of a signed distance map of the binary image. Let *B*(0) and *B*(*K*) be centrally located points on the opposite sides of the boundary such that 0 *< K < M*. *B*(0) and *B*(*K*) remain fixed and are assumed to be medially linked since the normals to the boundary at these points are approximately anti-parallel for thin oblong shapes. Let *q*_*U*_ and *q*_*L*_ be points on the interval [0, *M*) that map to opposites ends of the shape, such that *B* can be represented in terms of four quadrants: *U*_+_, *L*_+_, *L*_*−*_, *U*_*−*_. The superscripts (+, −) refer to symmetry about the medial axis, and (*U, L*) refer to symmetry about a line through *B*(0) and *B*(*K*). The optimal values of *q*_*U*_ and *q*_*L*_ are those for which *B*(*q*_*U*_) and *B*(*q*_*L*_) lie at the edges of the medial axis of the 3D shape and are not medially linked to any other points on *B*. As a result, medially linked boundary points on *B* encode a medial contour that approximates the ridge of the signed distance transform of the shape.

**Figure 3:**
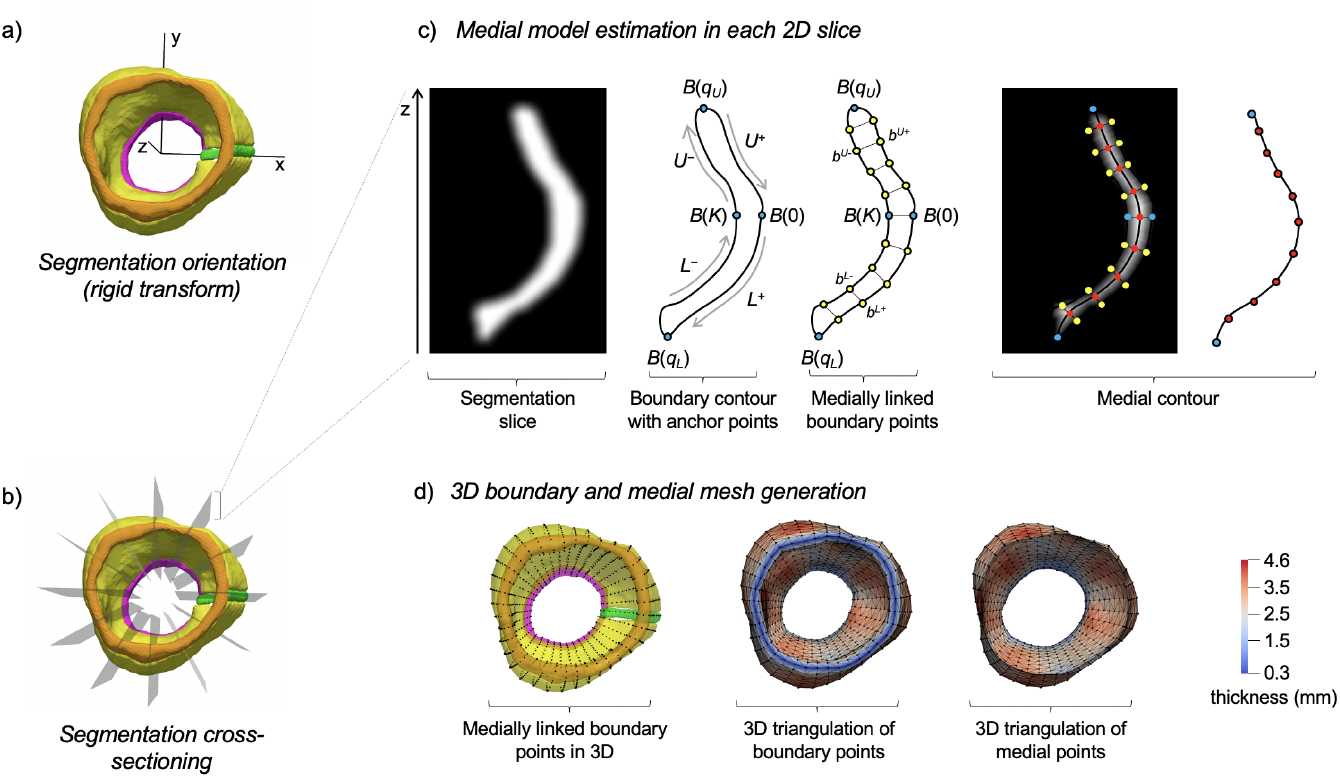
Generation of a 3D medial mesh from an image segmentation of the aortic root. (A)The aortic root segmentation is rigidly transformed so that the outflow tract is vertically oriented along the z-axis and the interatrial septum (green) is aligned with the x-axis. (B) The oriented segmentation is rotationally cross-sectioned into long-axis slices of the root. (C) In each cross-section, a 2D non-branching medial axis is approximated using a surface resampling technique, wherein points on the medial axis are estimated as midpoints between medially linked boundary points on the root surface. (D) 3D triangulated surfaces of the root boundary and medial surface are generated by defining edges between nodes in neighboring cross-sections. Locally varying thickness of the aortic root is shown in color.

For the aortic root, the values of *q*_*U*_ and *q*_*L*_ are computed from the vertical extrema of B, and the locations of medially linked boundary points *b* ∈ ℝ^2^ in each quadrant of *B* are subsequently calculated by contour interpolation as follows:

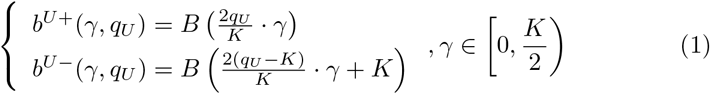

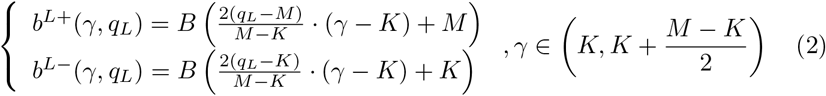

Note that in Equations 1 and 2, the medially linked boundary points need only be defined over the domain of two quadrants of *B* that are symmetric about a line through *B*(0) and *B*(*K*). Points on the medial axis **m** can then be estimated as the midpoints between pairs of medially linked boundary points. For thin cylindrical shapes like the aortic root, the estimated medial axis aligns well with the ridge of the signed distance transform shown in Figure 3C. For anatomies with larger shape variation, **m** can alternatively be updated through constrained optimization as described in [66], such that the values of *q*_*U*_ and *q*_*L*_ are optimized to align the medial axis with the ridge of the signed distance transform.

Once nodes on **m** are defined for each 2D cross-section of the shape, a 3D triangulated mesh is automatically created by defining edges between nodes of adjacent cross-sections. The edges define triangular faces such that the triangle normals point outward from the medial axis. Shown in color in Figure 3D, a scalar thickness measurement is associated with each node, which is defined as the distance between the medially linked boundary nodes from which the point on **m** was estimated. The resulting mesh is a medial model of the aortic root that has a standardized triangulation (i.e., consistent nodes, edges, and faces with fixed topology) regardless of the size of the input segmentation from which it is generated. Note that the procedure for medial mesh generation is only performed in one 3D frame (the frame in which the root was initially segmented in 3D). Thereafter, the same medial mesh is transformed to other 3D frames based using the affine transformations and deformation fields obtained in the propagation scheme described in Section 2.2.

### 2.4. Strain estimation

The medial model provides a spatial correspondence across different phases of the cardiac cycle since the 3D medial mesh is propagated in time using deformable registration, which facilitates strain calculation. The medial model also provides spatial correspondence between different patients since the segmentation-derived medial mesh is computed using a standardized boundary sampling scheme. In order to temporally align data for strain measurement, the first 3D TEE frame showing AV closure after systole was manually identified in each patient’s image series. The frame just prior to AV opening was considered to represent the lowest pressure and therefore the “mechanical reference configuration”.

Since the medial model is a two-dimensional manifold in three-dimensional space S ∈ ℝ^3^, it can be parameterized using coordinates *s*^*α*^ with *α* = 1, 2. If the spatial coordinates of the medial model nodes in the mechanical reference configuration are denoted as ***X***_*I*_, while those in other frames are denoted as ***x***_*I*_ (*t*). From the nodal coordinates, the entire surface is obtained by interpolation ***X*** = Σ_*I*_ *N*_*I*_ (*s*^*α*^)***X***_*I*_ and ***x***(*t*) = Σ_*I*_ *N*_*I*_ (*s*^*α*^)***x***_*I*_ (*t*). Piecewise linear Lagrangian polynomials [67] are used for *N*_*I*_ to calculate the in-surface strains, while *C*^2^-continuous Loop’s subdivision functions [68] are used to calculate the curvature strains. Once interpolated, the basis vectors are calculated as 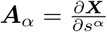 in the reference configuration and 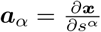 in the deformed configuration. From the reference covariant basis ***A***_*α*_, the components of the reference metric are calculated as *G*_*αβ*_ = ***A***_*α*_ · ***A***_*β*_ and the reference contravariant basis vectors are calculated using of the metric tensor as ***A***^*α*^ = [*G*_*αβ*_]^*−*1^ · ***A***_*β*_. Finally, the deformation gradient **F** = *∂****x****/∂****X*** is obtained by

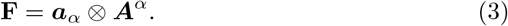

From **F**, the right Cauchy-Green deformation tensor is calculated as **C** = **F**^*T*^**F**, and its invariants are calculated as [69]

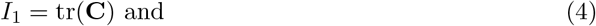

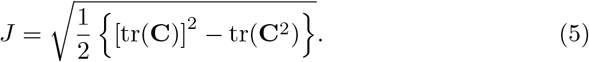

*Ī*_1_ = *I*_1_*/J* quantifies the shear strain (*Ī*_1_ ≥ 2 always and *Ī*_1_ = 2 represents zero shear strain) and *J* quantifies the area dilatational strain (*J* = 1 indicates no change in area). While it is common to decompose the in-plane strains in vessels into circumferential and longitudinal components, the chosen representation in terms of *Ī*_1_ and *J* offers two advantages. First, the representation in terms of invariants does not require circumferential and longitudinal direction vectors. Second, the two invariants represent different modes of deformation and, as a result, are much less correlated than other measures. *Ī*_1_ is termed as the shear strain and *J* is termed as areal or dilatational strain. In addition, the thickness obtained from medial model is converted into thickness strain, defined as the ratio of deformed thickness to the thickness at mechanical reference configuration.

The surface normals are calculated by using the cross product of basis vectors and normalizing it:

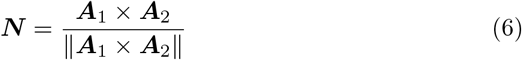

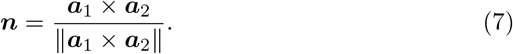

Curvature changes during the cardiac cycle can also be calculated (see Appendix) but, since no significant changes were found in curvatures, these are not reported in this study.

### 2.5. Quantitative and statistical analysis

Fitting the same medial mesh to all patients’ images provides a spatial correspondence. In order to determine the time correspondence, the frames when AV first opens and first closes were manually identified and marked. The images were assumed to be acquired at equally spaced time points and over a complete cardiac cycle. Therefore, in addition to the AV opening/closing time points, strain quantities at two additional time points – mid-systole and mid-diastole – were determined by piecewise interpolation and considered to correspond to the same time point for each patient.

Quantities defining the reference shape (assumed to be at one frame before the AV opens) were averaged across patients. The mean values of all strain-related quantities were calculated at the above four time points. To determine the statistical significance of differences between TAVs and BAVs, the non-parameteric Wilcoxon rank-sum test was carried out at each spatial point. However, no correction was applied for the multiple comparisons^2^.

### 2.6. Implementation

The methodology has been implemented as a publicly available distributed segmentation service in ITK-SNAP, entitled *GoValve-Root* at *dss*.*itksnap*.*org*. Documentation and an example are available at https://govalve-root-src.readthedocs.io/en/latest/ with source code at https://github.com/apouch/GoValve-Root-src.

### 2.7. Verification

In order to assess the accuracy of the tracking algorithm, each aortic root was manually traced again in a different cardiac phase such that there were two manual segmentations completed for each 4D image series. The propagation of the first manual segmentation was then evaluated with respect to the second manual segmentation in terms of the Dice overlap and symmetric distance metrics. For example, if a patient’s data set consisted of 20 frames and frame 3 was manually traced at systole, then another frame at diastole was randomly selected (e.g., frame 17) and manually traced. Then, the segmentation of frame 17 created by propagation of frame 3 was compared to the manual segmentation of frame 17. This evaluation was carried out for each patient in the study.

In order to verify the accuracy of strain estimation, three TAV and three BAV manually TEE images were chosen at random and their manually segmented frames were used as the starting (reference) image. An affine, isotropic deformation of a maximum of 10% stretch was applied, based on expected maximum stretch in-vivo. The affine transformation was applied in 10 incremental steps to create artificial 4D datasets with 10 time frames each, where the true strains were known a priori based on the applied deformation. A percentage error between the true strain values and estimated strain values were calculated for the three types of strains – dilatational strain, shear strain, and thickness strain.

## 3. Results

### 3.1. Reference geometry

Comparison of aortic root reference geometry in the TAV and BAV groups is shown in Fig. 4. The average TAV and BAV root shapes are shown in Fig. 4a with locally varying luminal radius in color. In order to better visualize locally resolved measurements such as luminal radius, strain, and thickness, the measurements are illustrated on a disk-shaped projection of the 3D root onto a plane perpendicular to the outflow tract. In this mapping (e.g., Fig. 4d), the inner contour of the disk-shaped projection represents the LVO and the outer contour represents the STJ. A similar representation, known as the bulls-eye plot is used in cardiac research [70]. While the TAVs and BAVs analyzed in this study fall within a designated size range, the plots of luminal radius show greater prominence and consistency in the sinuses of Valsalva in TAV roots than in BAV roots. The average luminal radius and ellipticity along the length of the root are compared in Fig. 4e,f. While there is no significant difference in the luminal radius except near the STJ (as the data sets in this study were intentionally selected with respect to root size), the sinuses of TAV roots have a higher ellipticity. Thus, even when matched for size, there are observable differences in TAV vs. BAV root shape (Fig. 4g), especially at the sinuses of Valsalva. Differences in root wall thickness were also observed, with BAV roots being thinner than the TAV roots, especially at the sinuses (Fig. 5). This difference in thickness can have an important effect on the in-plane and bending stiffnesses of the arterial tissue.

**Figure 4:**
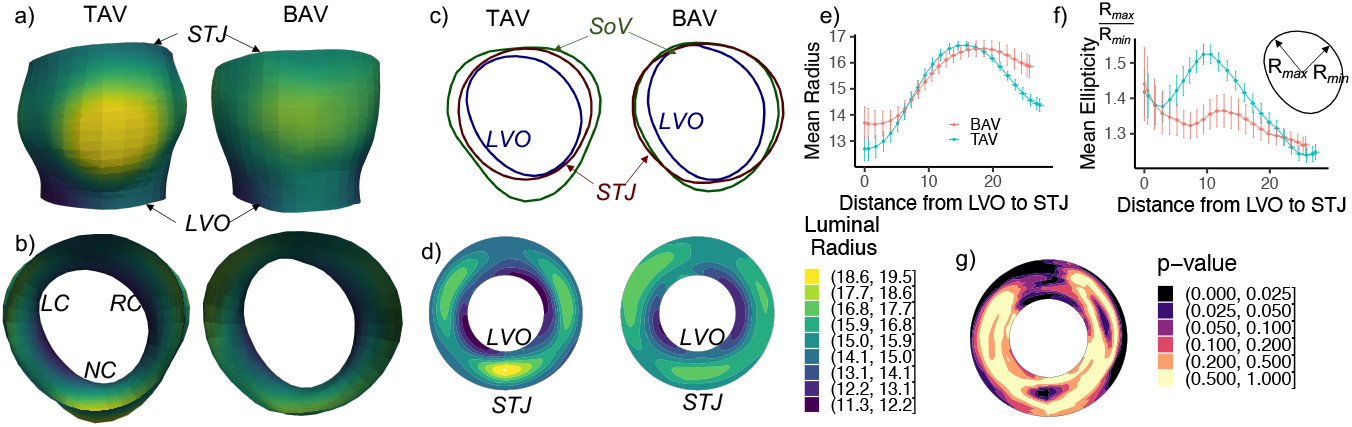
a,b) The average reference shape of aortic root for patients with TAV versus BAV in two orientations colored by luminal radius and indicating the locations of left coronary (LC), right coronary (RC), and non-coronary (NC) sinuses. c) Three slices of the reference shape showing the LVO, sinus of valva (SoV), and STJ. d) A polar contour plot of the luminal radius where inner radius is the LVO and outer radius is the STJ. e) Radius and f) ellipticity of the reference shapes along the length of the root. g) Polar contour plot of the p-values comparing the luminal radius of TAV and BAV roots.

**Figure 5:**
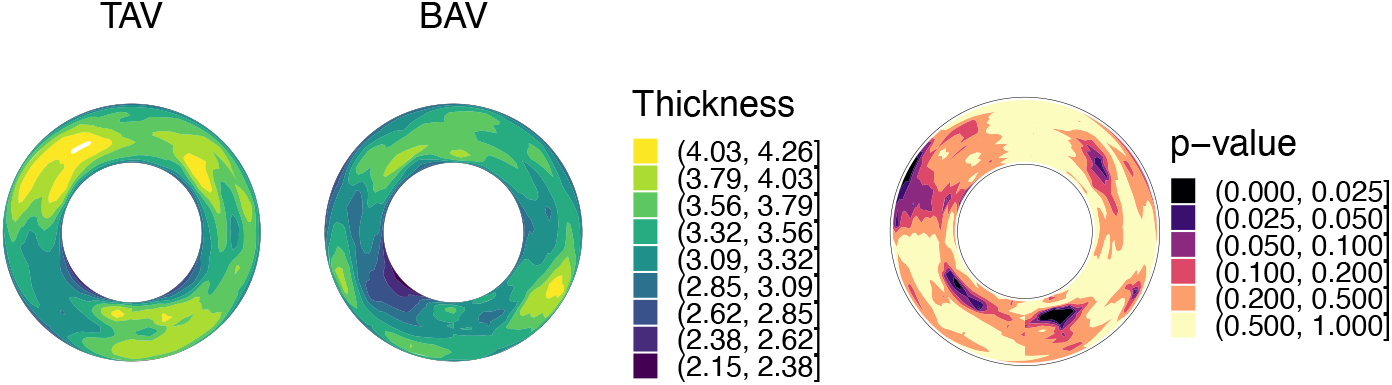
Average thickness of the root wall at the reference configuration for patients with TAV and BAV, and the p-value of differences between them tested using Wilcoxon rank-sum test.

### 3.2. Assessment of tracking accuracy

The Dice overlap between the propagated and manual segmentations for the 17 data sets averaged 0.72 ± 0.07. The symmetric mean boundary distance averaged 0.31 ± 0.11 mm.

### 3.3. Verification of estimated strains

A boxplot of percentage errors in the three estimated strains for six (there TAV and three BAV) images are shown in Fig. 6. The errors were negligible in the shear strain since no shear was induced when applying an affine transformation. Moreover, 95 percentile of the errors in the dilatational and thickness strains were less than 5%. As the deformation level increases, the ranges of errors increased slightly.

**Figure 6:**
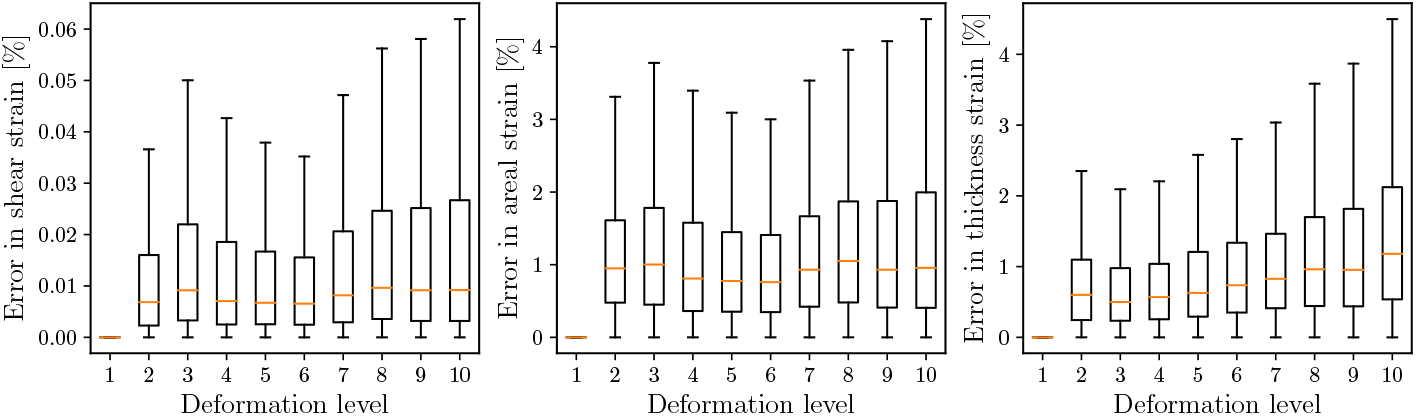
The three strain estimates (shear strain, dilatational strain and thickness strain) for six artificial datasets show an acceptable error range.

### 3.4. Strains over the cardiac cycle

Since strains depend on the gradient of the deformation, they require a choice of “reference” configuration, with respect to which the gradients are calculated. To this end, the time frame when the AV first opens was identified manually, and one frame before that was chosen as the reference configuration. This assumption is based on the general rule that pressure in the aortic root is minimum just before AV opening.

Both global and regional strains were computed from the image-derived 4D models of the aortic root. Global strain is a measure of change in the total area/volume of the root during the cardiac cycle. To assess global strain, the total root wall surface area, wall volume (defined as the sum of wall area multiplied by wall thickness) and the lumen volume enclosed by the root were calculated at all time points, and their ratios with respect to the reference configuration were plotted (Fig. 7). No statistically significant differences were observed with respect to these global measures in the TAV and BAV roots analyzed in this study.

**Figure 7:**
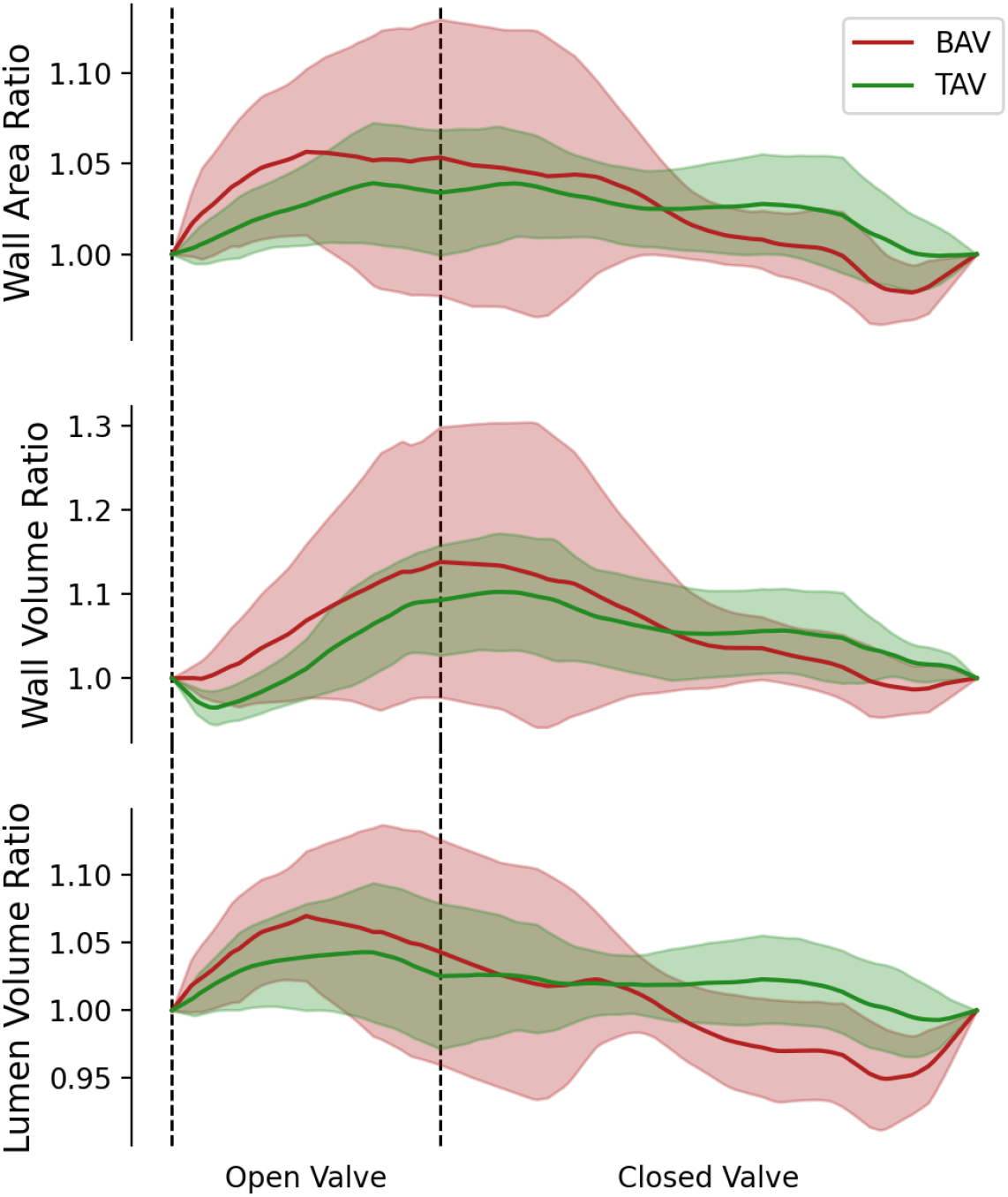
The global strains quantified in terms of the ratio of aortic root wall area, aortic root wall volume, and aortic root lumen volume for patients with TAV and BAV.

Regional strains were calculated and quantified in terms of shear and dilatational strains (*Ī*_1_ and *J*), with the mean values at four time points plotted in Fig. 8a,b. Differences in regional strain in BAV versus TAV roots were observed, with regional strains of BAV roots being higher. Similarly, the strain in wall thickness was calculated and its mean values are graphed in Fig. 8c, which also shows higher values in BAV roots.

**Figure 8:**
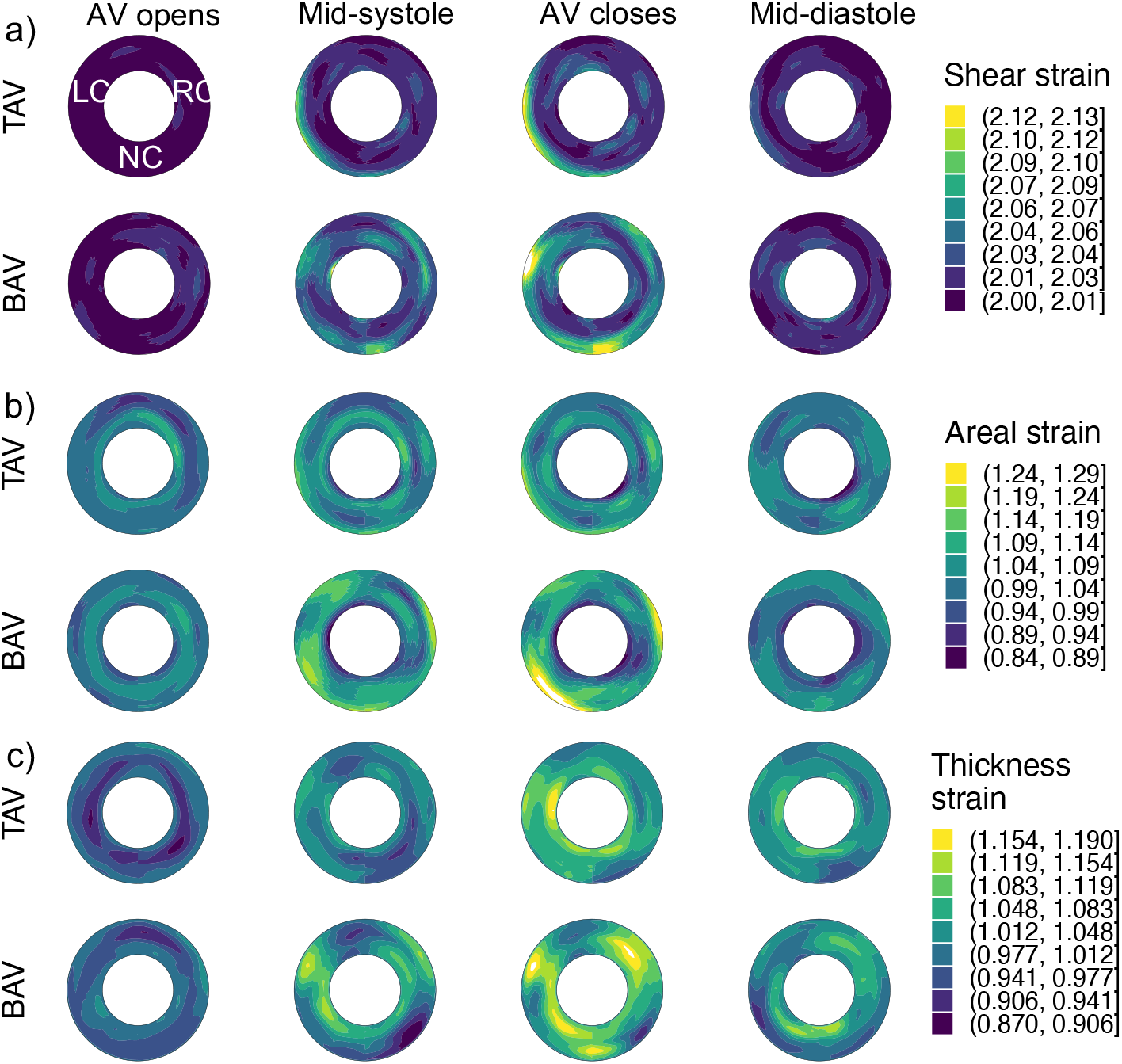
The average strains in aortic root for patients with TAV versus BAV at four points in the cardiac cycle: a) shear strain, b) areal strain, and c) thickness strain.

The results from Wilcoxon test comparing the strains at all four time points between TAVs and BAVs were plotted (Fig. 9). These plots confirm that there are statistically significant differences between the regional strains, with aortic roots in BAV patients experiencing higher strains. Interestingly, these differences are in slightly different areas of the root as well. Higher shear strains are observed in the region where the left ventricular outflow jet is directed towards the aortic arch, which could be related to the differences in the hemodynamics in TAVs and BAVs. No significant differences were found in the bending strains, quantified by changes in the curvature (result not shown here for brevity).

**Figure 9:**
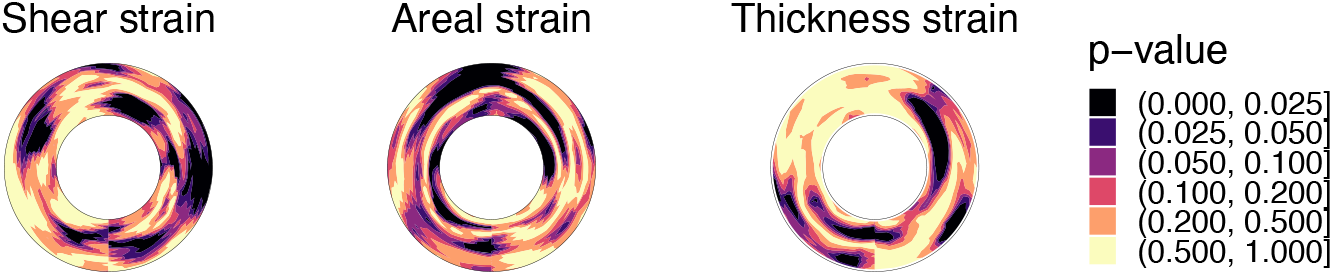
Differences between the measured strains in the aortic root for patients with TAV versus BAV tested using Wilcoxon rank-sum test

## 4. Discussion

This study presents a novel approach to estimating 3D tissue strains invivo in thin tubular structures, for which the aortic vasculature is a clinically motivated example. Development of this approach is a necessary first step towards investigating whether characterization of locally resolved deformations can contribute to our understanding of the pathophysiology of aortopathies, as well as the prediction of their progression. The method combines deformable registration and medial modeling with large deformation kinematic calculations commonly used in nonlinear mechanics. While this work demonstrates application to the mechanics of the aortic root, the proposed methodology is general and can be applied to any structure with tubular topology for which 4D images are available. The application to the aortic root, in particular, is clinically motivated since BAVs are commonly associated with progressive pathologies not only of the cusps (e.g., stenosis and regurgitation), but of the aortic root and ascending aorta as well [71]. An open-source example of the pipeline has been made available through ITK-SNAP’s distributed segmentation service for further research applications.

In addition to presenting a novel pipeline for image-to-strain analysis, a goal of this study is to demonstrate a clinical application to anatomical shape and tissue mechanics assessment that can be carried out with the proposed algorithm. In terms of anatomical form, BAV patients commonly have larger aortic roots than patients with TAVs. To minimize the effect of root size on strain analysis, TAV and BAV cases with comparable root size were selected for this study. Nevertheless, in these cases with similar root dimensions, differences were observed in the local root morphology. Specifically, the sinuses of Valsalva were observed to be more pronounced in TAV patients, likely owing to the consistency of cusp and sinus morphology in physiologically normal TAVs. The luminal radius at the LVO and STJ were slightly larger in the BAV roots, which is consistent with frequent clinical observation of STJ dilatation in the BAV population. Moreover, the root wall thickness in BAV patients was found to be lower, which may affect effective in-plane and bending stiffnesses of the tissue.

Global aortic root strains were not found to be significantly different in the BAV and TAV groups in this study, thus motivating the need for localized strain assessment. In terms of regional strain, there was a clear difference between the TAV and BAV groups, with BAV patients experiencing higher shear, dilatational, and thickness strains in certain areas of the root. Interestingly, the between-group differences in these three strain measures localized to different regions of the root. For example, shear strains were found to be higher near the STJ above the right coronary (RC) sinus, the dilatational strains were higher at the LVO and the STJ in the region between the left and right coronary sinuses. Lastly, there was a small area near the non-coronary (NC) and right coronary (RC) sinuses, where the thickness strains were higher in the BAV. These results show the significance of strain analysis, which may shed light on the role of tissue mechanics in aortic root pathology when applied to larger populations. In addition, future longitudinal studies could investigate whether strains could be leveraged as a prognostic index related to risk of aortic dilatation, dissection, or rupture. To allow for application to future studies, the proposed framework has been made openly available as a distributed segmentation service (DSS) in ITK-SNAP.

While this work demonstrates a novel, generalizable shape and strain analysis pipeline, there are a few limitations of the approach. First, the analysis used manual segmentation of the root in one 3D volume of each 4D data set, which will be automated in future work. Second, the choice of reference configuration for strain calculation has limitations, as there is no point in the cardiac cycle when aortic tissue is truly load-free. The choice of the mechanical reference frame in this study was based on a commonly accepted point of lowest pressure in the cardiac cycle. As a result, the resulting strains should be viewed with respect to the diastole and not with respect to the true stress-free state (which would require tissue explantation and is not possible in-vivo). Thus, these strain estimates with respect to diastole do not provide a complete picture of the biomechanical state. Third, the accuracy of the calculated strains depends on the image registration algorithm. Since image registration is well-known to be an ill-conditioned problem, the results may be influenced by regularization, which is a limitation of all image-based strain analysis assessment tools. Thus, the same regularization parameters were used for all datasets in the study and we consider the reported strains to be estimates, which could potentially be improved with physics-inspired regularization or by analysis of the raw signal. Since commercial echocardiography machines generally do not output raw data, the effect of different regularization approaches will be a topic of future studies. Fourth, future work will specifically investigate the reproducibility of thickness strain estimation with respect to another imaging modality such as CT. Since the root wall is relatively thin (only several voxels thick in 3D TEE), thickness strain is more likely impacted by image resolution than areal and shear strains. Finally, the population of TAVs and BAVs analyzed in this study was relatively small, so the goal of reporting between-group differences in root mechanics is intended to illustrate the clinical potential for application of the image-to-strain analysis pipeline methodology to larger populations, rather than to draw conclusions about BAV root pathophysiology.

In the future, the framework will be adapted to other, more complex topologies and can be used towards estimating mechanical stresses as well. For different structures with non-cylindrical morphology, new medial modeling methods will be adapted. For thick structures, such as myocardium, a three-dimensional kinematics (as opposed to the proposed shell kinematics) will be implemented. Calculation of stresses will require choosing a stress-strain relationship and the associated parameters, as well as the temporal and spatial variations of the luminal pressure, adding further complexity to the problem. However, it might be possible to treat these properties stochastically and approximate their distributions from the tissue deformation through the cardiac cycle.

In conclusion, this work presents a semi-automated methodology for estimating global and regional strains in-vivo in a manner that enables statistical comparison between patients. The method is based on fully nonlinear shell-based kinematics, which divides the strains into in-plane (shear and dilatational) and out-of-plane. The results suggest that relative to size-matched non-aneurysmal TAV roots, BAV patients experience higher regional shear strains in their aortic roots. This difference in strains might be a contributing factor to their higher risk of aortic root dilatation. The proposed framework is openly available and applicable to any tubular structures.

## Data Availability

All data produced in the present study are available upon reasonable request to the authors

## A. Appendix

The curvature tensor in reference and deformed configurations are calculated using the second derivative of interpolated positions as

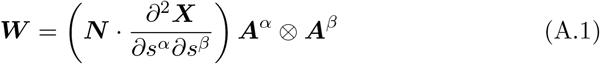

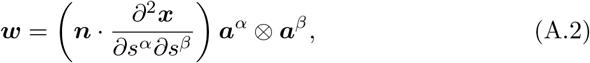

respectively. Mean curvature in the reference configuration is 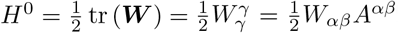. Similarly, the mean curvature in the current configuration is 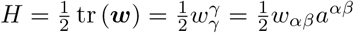. The Gaussian curvature is the determinant of curvature tensor 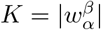.

## Footnotes

1 Luminal radius averaged circumferentially at the reference configuration

2 Due to strong spatial correlation, Bonferroni correction was considered to be too conservative.

